# Adequacy of serial self-performed SARS-CoV-2 rapid antigen-detection testing for longitudinal mass screening in the workplace

**DOI:** 10.1101/2022.02.10.22270805

**Authors:** Jesse Papenburg, Jonathon R. Campbell, Chelsea Caya, Cynthia Dion, Rachel Corsini, Matthew P. Cheng, Dick Menzies, Cedric P Yansouni

## Abstract

**Importance:** Longitudinal mass testing using rapid antigen detection tests (RADT) for serial screening of asymptomatic persons has been proposed for preventing SARS-CoV-2 community transmission. The feasibility of this strategy relies on implementation of accurate self-performed RADT testing where people live, work, or attend school.

**Objective:** To quantify the adequacy of serial self-performed SARS-CoV-2 RADT testing in the workplace, in terms of the frequency of correct execution of procedural steps and accurate interpretation of the range of possible RADT results. We compared results using the instructions provided by the manufacturer to those with modified instructions that were informed by the most frequent or most critical errors we observed.

**Design:** Repeated cross-sectional, diagnostic accuracy study performed prospectively in the field.

**Setting:** Businesses in Montreal, Quebec, Canada, with at least 2 active cases of SARS-CoV-2 infection.

**Participants:** Untrained, asymptomatic persons in their workplace, not meeting Public Health quarantine criteria.

**Exposures:** A Modified Quick Reference Guide compared to the original manufacturer’s instructions.

**Main Outcome(s) and Measure(s):** The difference in the proportions of correctly performed procedural steps, and the difference in proportions of correctly interpreted RADT proficiency panel results. The secondary outcome, among subjects with two self-testing visits, compared the second to the first self-test visit using the same measures.

**Results:** Overall, 1892 tests were performed among 647 subjects. For self-test visit 1, significantly better accuracy in test interpretation was observed using the Modified Quick Reference Guide for weak positive (55.6% vs. 12.3%; 43.3 percentage point improvement, 95% confidence interval [CI] 33.0%-53.8%), positive (89.6% vs. 51.5%; 38.1% difference, 95%CI 28.5%-47.5%), strong positive (95.6% vs. 84.0%; 11.6% improvement, 95%CI 6.8%-16.3%) and invalid (87.3% vs. 77.3%; 10.0% improvement, 95%CI 3.8%-16.3%) tests. Use of the modified guide was associated with smaller, statistically significant, improvements on self-test visit 2. For procedural steps identified as critical for the validity of test results, adherence to procedural testing steps did not differ meaningfully according to instructions provided or reader experience.

**Conclusions and Relevance:** Longitudinal mass RADT testing for SARS-CoV-2 can be accurately self-performed in an intended-use setting; this work provides evidence for how to optimise performance.

**Key Points:** *Questio:* Do untrained users correctly perform and interpret the results of SARS-CoV-2 rapid antigen detection tests (RADT) in the workplace, and how can their performance be optimised?

*Findings:* In this prospective field evaluation of self-performed SARS-CoV-2 RADT in an intended-use setting, we found that the accuracy of RADT interpretation was poor when the manufacturer’s instructions were used. A Modified Quick Reference Guide yielded significantly better user performance.

*Meaning:* Longitudinal mass RADT testing for SARS-CoV-2 can be accurately self-performed in an intended-use setting; this work provides evidence for how to optimise performance.

## INTRODUCTION

Ongoing transmission of SARS–CoV-2^1^ is frequently driven by asymptomatic or pre-symptomatic individuals.^2^ Mass deployment of lateral flow rapid antigen detection tests (RADT) for serial screening of asymptomatic persons has been proposed for preventing community transmission, with the aim of promptly detecting individuals most likely to be infectious.^3,4,5,6^ This strategy may be even more relevant with the emergence of the Omicron (B.1.1.529) variant^7^ that may evade immunity afforded by vaccination.

The feasibility of longitudinal mass testing relies on implementation of self-performed RADT where people live, work, or attend school. However, several factors impede this deployment. These include regulatory requirements that RADT be performed by trained health care workers in some countries^8^ and lack of data on the accuracy of self-administration in their intended-use settings.^9^ Similarly, no studies have assessed the impact of repeated lateral flow testing on user performance.

We report the results of a prospective field evaluation of the accuracy of serial self-performed RADT for SARS-CoV-2 among untrained, asymptomatic persons in their workplace.

## METHODS

### Study design and setting

In this repeated cross-sectional, diagnostic accuracy study, businesses in Montreal, Quebec, Canada, with at least 2 active cases of SARS-CoV-2 infection within a 14 day period were identified by the Montreal Department of Public Health. For businesses that agreed to participate, study visits were scheduled twice weekly for two weeks (4 visits). Study data were collected and managed using REDCap.^10^

Informed consent was obtained for each participant. The study was approved by the Research Ethics Board of the Research Institute of the McGill University Health Centre (MP-37-2022-7762).

### Diagnostic testing procedures

The RADT used was the Panbio COVID-19 Ag Rapid Test Device (Abbott Laboratories, Saint-Laurent, Quebec). At each visit, trained study personnel instructed participants on procedures to perform self-collected nasal mid-turbinate swab specimens. On visits 1 and 3, all post-specimen collection steps and result interpretations were performed by study personnel. On visits 2 (first self-testing visit) and 4 (second self-testing visit), these steps were performed by the participant. During self-testing, study personnel assessed 13 RADT procedural steps performed by the participant as correct or incorrect. If any of 6 steps deemed to be critical to testing integrity were assessed as incorrectly performed, the participant was informed at the end of the procedure and testing was redone by study personnel. Participants with a positive RADT result were offered confirmatory laboratory-based PCR testing at a local testing centre.

### Result interpretation using a proficiency panel

A proficiency panel of 7 RADT test results was created using serial dilutions of the positive control reagent and aimed to evaluate participants’ ability to interpret the range of possible signal intensities of the test line, including “weak positive”, “positive”, “strong positive”, “invalid” and “negative” test results (eFigure 1). The panel was printed in colour at actual size and high resolution on cardboard. At visits 2 and 4 participants were asked to interpret the 7 RADT results, in addition to their own RADT result.

### Intervention

Participants were initially given access to the complete manufacturer’s instructions, the visual Quick Reference Guide (eFigure 2), and no time limit during self-testing visits. Study personnel did not give any additional guidance. Following a planned interim analysis, we developed a Modified Quick Reference Guide (eFigure 3) that addressed the most frequently observed user errors in RADT procedural steps and interpretation, and deployed it subsequently.

### Outcomes

Among all subjects with at least one self-testing visit, we compared the group using our Modified Quick Reference Guide to the group using the original manufacturer’s instructions. Among subjects with two self-testing visits, we compared the second to the first self-test visit. For each comparison, our primary outcome was the difference in the proportion of correctly interpreted RADT proficiency panel results. Our secondary outcome was the difference in proportion of correctly performed procedural steps.

Information on data collection, sample size estimate, and statistical analysis are provided in Supplementary Material.

## RESULTS

From July 7 to October 8, 2021, 236 outbreak businesses were identified (Figure 1); 168 (71%) were contacted and 13 (5%) participated. Overall, 1892 tests were performed among 647 subjects (mean of 2.9 visits per subject), of which 278 had at least 1 self-testing visit (115 with the Modified Quick Reference Guide; eTable 1). RADT result was positive in 3/1892 (0.16%), of which 1 was confirmed positive and 2 were negative by PCR.

**FIGURE 1:**
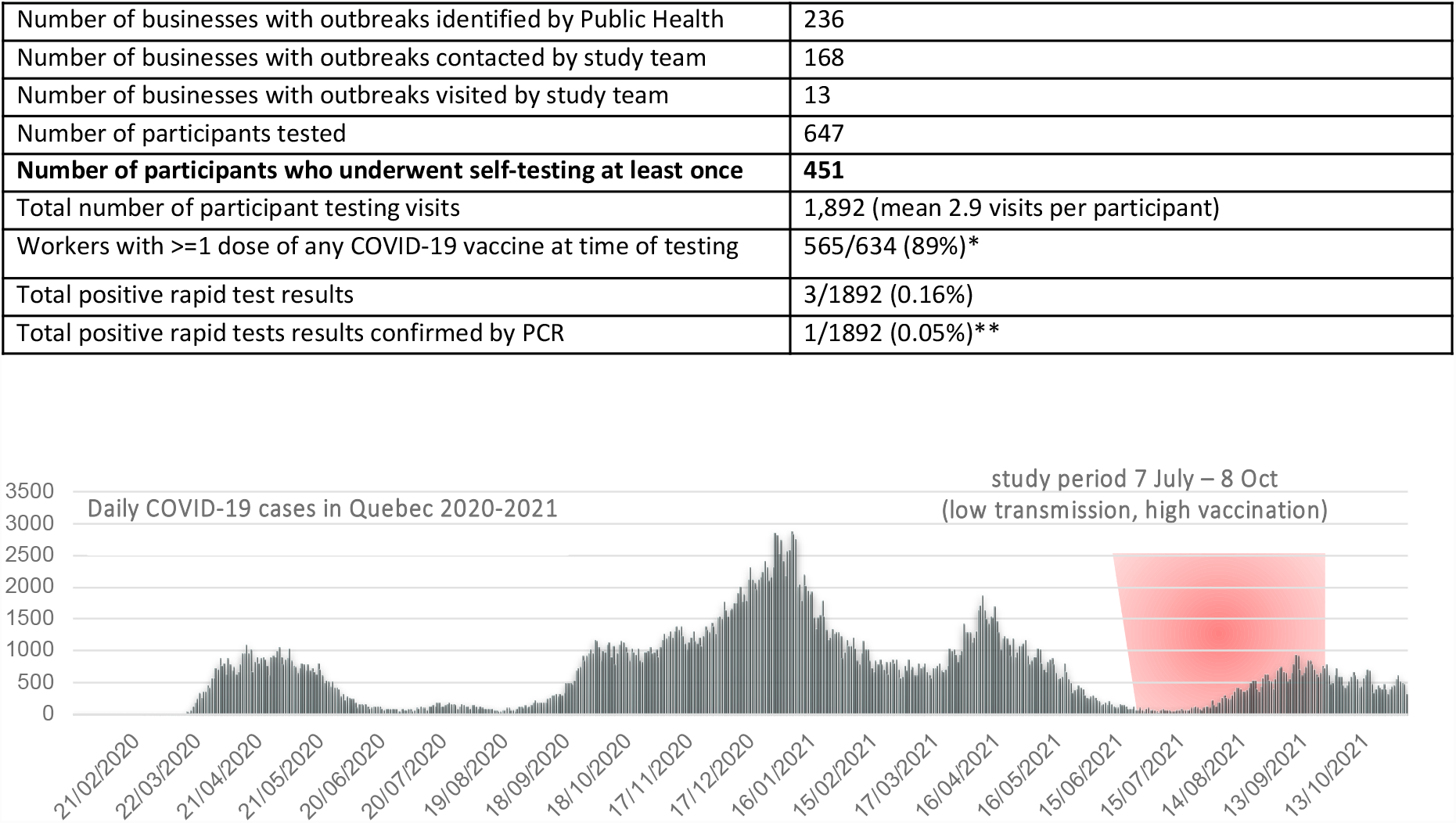
Summary of study recruitment and epidemiologic context. * Among the 634 of 647 participants who answered the question about vaccination status ** All positive RDT results underwent confirmatory PCR testing on the same day according to the study protocol. Data on daily COVID-19 cases available at https://www.donneesquebec.ca/recherche/dataset/covid-19-portrait-quotidien-des-cas-confirmes. Modified Quick Reference Guide implemented as of Sept 13, 2021.

At the first-self-test visit, significantly better accuracy in interpreting proficiency panel RADT results was observed using the Modified Quick Reference Guide for weak positive (55.6% vs. 12.3%; 43.3 percentage point improvement, 95%CI 33.0%-53.8%), positive (89.6% vs. 51.5%; 38.1% improvement, 95%CI 28.5%-47.5%), strong positive (95.6% vs. 84.0%; 11.6% improvement, 95%CI 6.8%-16.3%) and invalid (87.3% vs. 77.3%; 10.0% improvement, 95%CI 3.8%-16.3%) tests. Use of the modified guide was associated with smaller, yet statistically significant, improvements on self-test visit 2 for weak positive, positive and invalid samples (Figure 2). In multivariable models accounting for outbreak business, participant age and gender, the Modified Quick Reference Guide was associated with higher accuracy in result interpretation for self-test visit 1 (adjusted odds ratio [aOR] 13.66; 95%CI 4.93-37.84), but not for self-test visit 2 (aOR 0.86, 95%CI 0.13-5.51).

**FIGURE 2:**
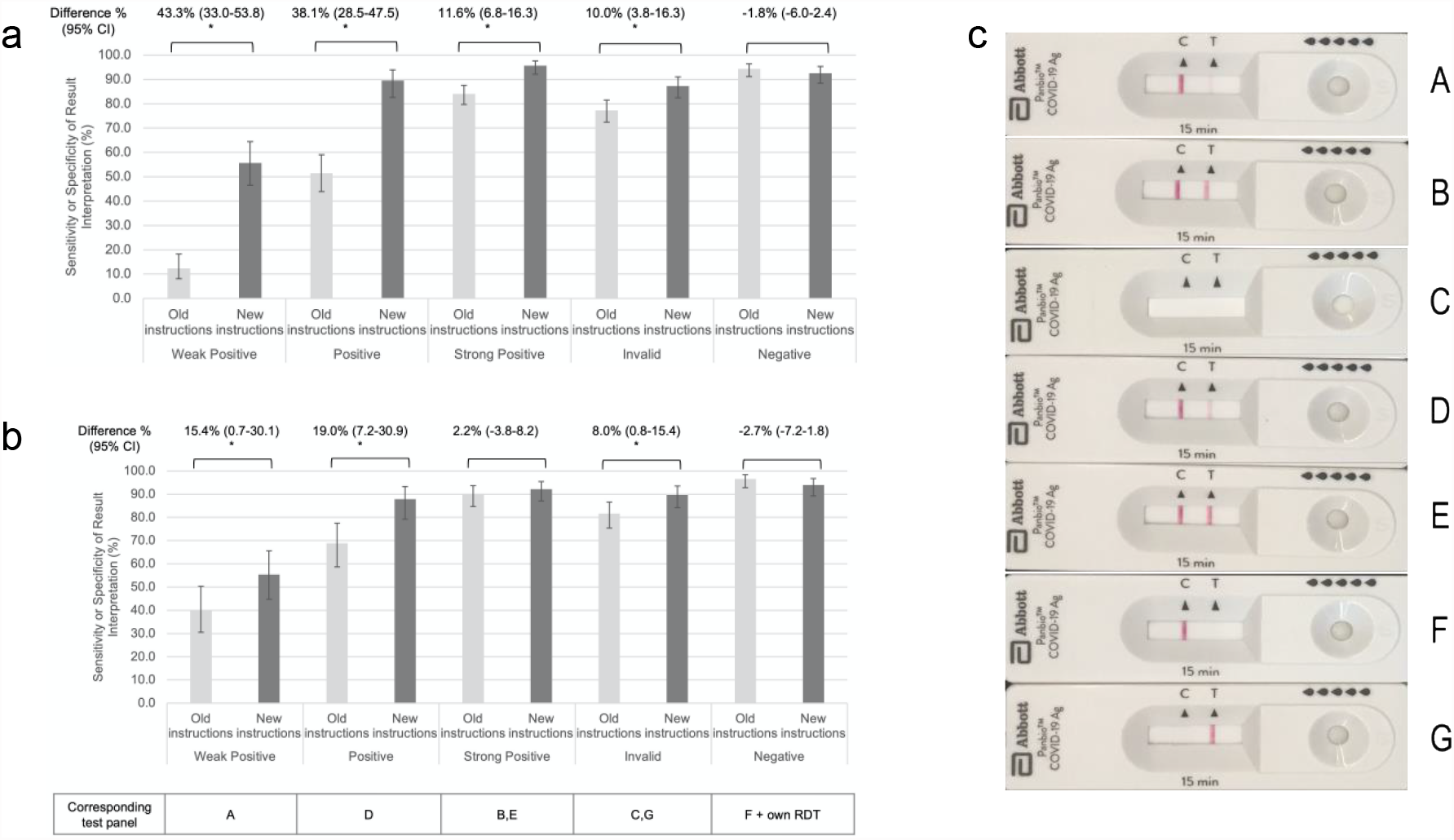
Reader accuracy for each type of RDT result interpretation, according to instructions provided and reader experience. Light grey bars show reader accuracy among participants undergoing self-testing using the instructions and quick reference guide provided by the manufacturer. Dark grey bars show reader accuracy among participants undergoing self-testing using the Modified Quick Reference guide designed by the investigators to address the most frequently observed errors. Panel a shows reader accuracy among those with a first self-testing visit (n=278 of whom 163 used the older version of the instructions provided by the manufacturer and 115 used the modified version). Panel b shows reader accuracy among those attending a second self-testing visit (n=173 of whom 90 used the older version of the instructions and 83 used the modified version). Panel c shows the panel of RDT results used to assess reader accuracy. Error bars represent the 95% confidence intervals for the sensitivity or specificity of the result interpretation estimated according to a binomial distribution using the Wilson Score method. Sensitivity refers to the accurate RDT result interpretations of positive and invalid results while specificity refers to the accurate RDT result interpretation of negative results. Differences in the estimated sensitivity/specificity between new and old instructions with 95% confidence intervals are also presented.

For procedural steps identified as critical for the validity of test results, adherence did not differ meaningfully according to instructions provided or reader experience (Figure 3).

**FIGURE 3:**
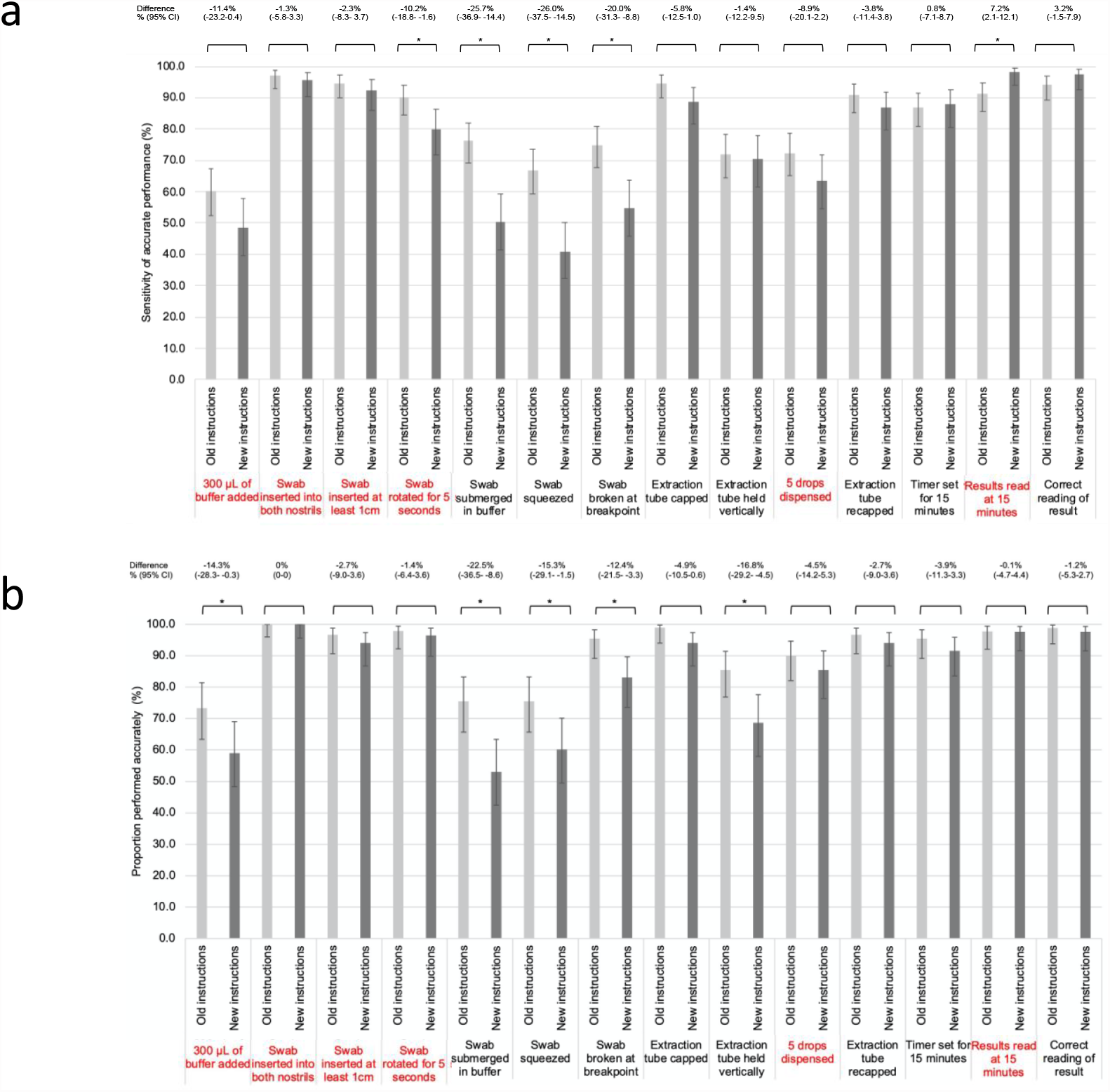
Adherence to procedural testing steps, according to instructions provided and reader experience. Light grey bars show adherence to procedural steps among participants undergoing self-testing using the instructions and quick reference guide provided by the manufacturer. Dark grey bars show adherence to procedural steps among participants undergoing self-testing using the Modified Quick Reference guide designed by the investigators to address the most frequently observed errors. Panel a shows adherence to procedural steps among those with a first self-testing visit (n=278). Panel b shows adherence to procedural steps among those attending a second self-testing visit (n=173). Steps in highlighted in red were those deemed to be most important for a valid result according to literature or expert opinion. Error bars represent the 95% confidence intervals for the proportion of participants who performed each step correctly, estimated according to a binomial distribution using the Wilson Score method. Differences between new and old instructions with 95% confidence intervals are also presented.

## DISCUSSION

In this prospective field evaluation of self-performed SARS-CoV-2 RADT in an intended-use setting, the accuracy of signal interpretation was proportional to test band intensity, and was extremely low for tests with weak bands. An evidence-based Modified Quick Reference Guide improved signal interpretation by untrained workers to a level of accuracy comparable to that of professional technologists for other RADTs,^11,12^ but did not have a major impact on the proper execution of procedural steps. Repeated self-testing was associated with improved signal interpretation, though the effect was less apparent with tailored instructions as initial accuracy was already high.

Self-testing using RADT has been studied for other infections. Among users attending a pre-travel clinic, 70.6% of all malaria RADT strips were correctly interpreted by untrained users.^13^ In studies of malaria RADT, weak positive test bands consistently correlate with lower sensitivity.^11–13^ Our study confirms these findings for SARS-CoV-2 RADT, and provides evidence for a simple and effective corrective intervention.

Study strengths include the recruitment of participants from diverse ethnic, socioeconomic and educational backgrounds. The Modified Quick Reference Guide was informed by the most frequent or most critical errors observed and validated with different participants. Further, we assessed the independent effects of modified instructions and repeated self-testing. Limitations include the small proportion of eligible businesses that entered the study, and that a single RADT was assessed with uncertain generalizability to the full diversity of available RADTs.

Self-administered RADTs are inexpensive and can be decentralised and implemented at scale for serial mass testing of asymptomatic persons to break chains of transmission and reduce SARS-CoV-2 incidence.^14,15^ This work shows that longitudinal mass SARS-CoV-2 RADT testing can be accurately self-performed and provides evidence for optimizing performance. This use case may become more pertinent with the emergence of new SARS-CoV-2 variants of concern.

## Supporting information

Supplemental Content

## Data Availability

All data produced in the present work are contained in the manuscript.

## Acknowledgments

The authors wish to thank the *Santé au Travail* Group of the Montreal Public Health Department, the office of the Mayor of the Borough of Montréal Nord, and the business owners and employees who participated in the study.

