## Supplemental Content for "Adequacy of serial self-performed SARS-CoV-2 rapid antigen-detection testing for longitudinal mass screening in the workplace"

**Supplemental Online Content**

### **eFigure 1: Serial dilutions of the positive control reagent and corresponding color scale.**

### **eFigure 2: Manufacturer’s Quick Reference Guide.**

### **eFigure 3: Modified Intervention Instructions.**

### **eTable 1. Instruction type used at self-testing visits 1 and 2.**

### **eMethods**

### **eFigure 1: Serial dilutions of the positive control reagent and corresponding color scale.**

Alignment of the positive tests used to generate the evaluation panel in the present study, against a standard colour scale. The rapid diagnostic test tested span a broad range of possible signal intensities of the test line. The test line is shown in the dashed rectangle.

### **eFigure 2: Manufacturer’s Quick Reference Guide.**

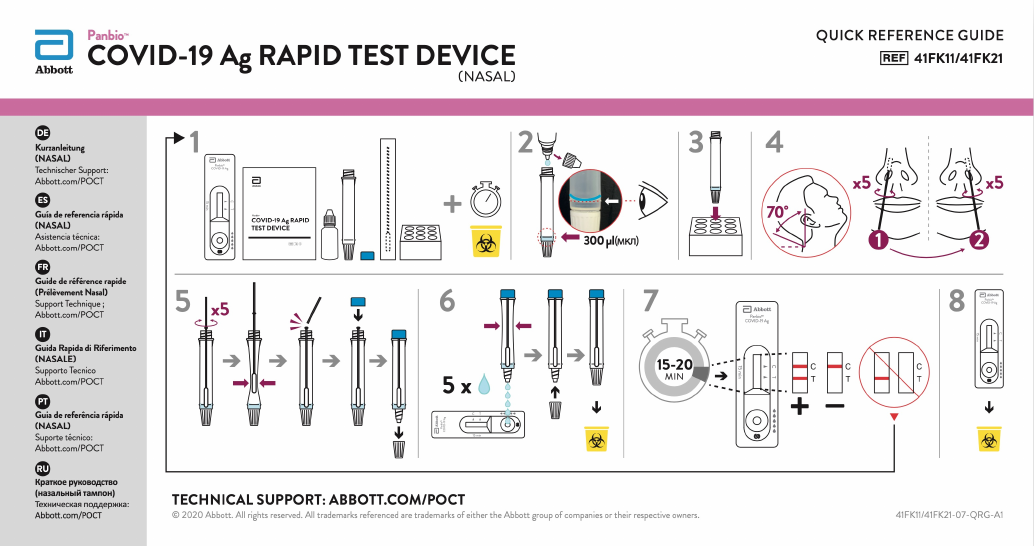

### **eFigure 3: Modified Intervention Instructions.**

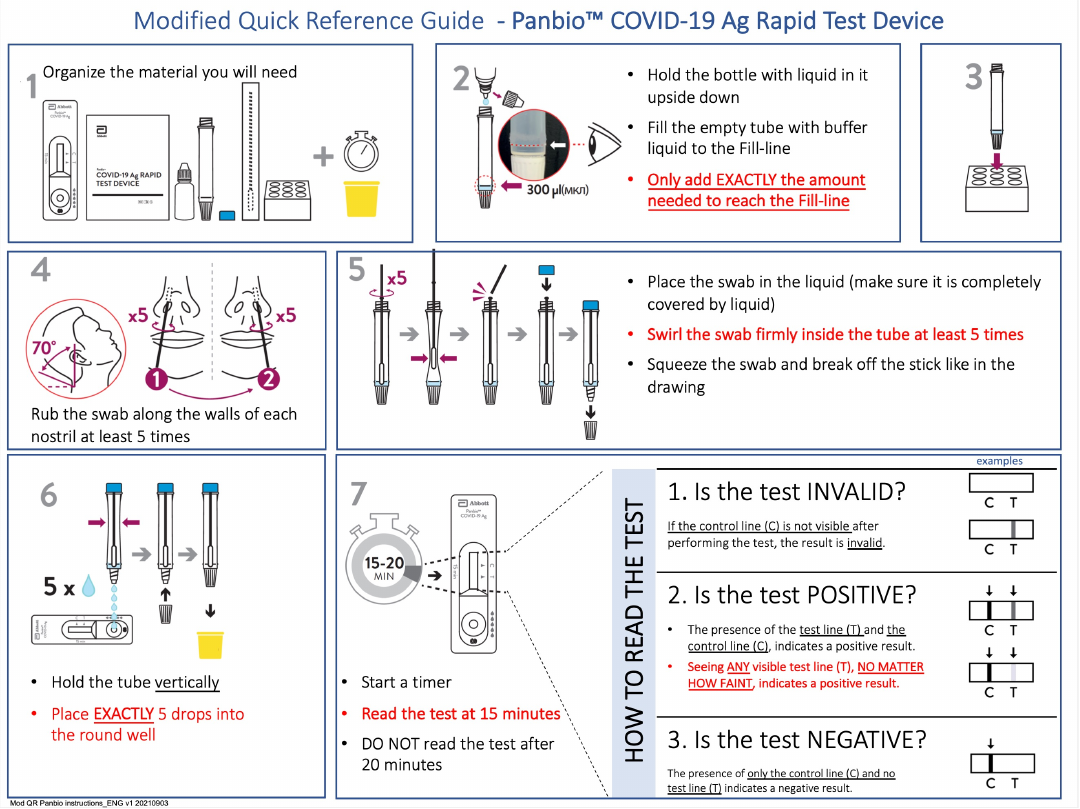

### **eTable 1.** Instruction type used at self-testing visits 1 and 2.

|  | **Self-testing visit 1**  **n=278** | **Self-testing visit 2**  **n=173** | **Total**  **n=451** |
| --- | --- | --- | --- |
| Manufacturer’s instructions used | 163 | 90 | 253 |
| Modified Quick Reference Guide used | 115 | 83 | 198 |

The modified quick reference guide was implemented as of Sept 13, 2021.

### **eMethods**

**Data collection**

Data on participant age, gender, language preference (English or French), COVID-19 symptoms, exposures, vaccination, as well as testing result and adequacy of performance of test procedures and result interpretation were collected.

**Sample size estimate**

A Modified Quick Reference Guide informed by the analysis of phase 1 data was expected to yield at least a 12% improvement in RADT reading accuracy over the manufacturer's instructions. Based on an expected overall weighted average of the accurate readings of 60% (95% CI 54-66) in phase 1, the number of readings required to achieve an accuracy of 72% with 6% precision was 216 readings per result category (invalid, negative, positive-weak, positive-medium, positive-strong). At the rate of two readings per person per visit, this corresponds to a recruitment of 108 people using the new instructions.

**Statistical analysis**

Baseline characteristics were summarized according to self-testing visit and reference guide type. Estimates of 95% confidence intervals (95% CI) around a proportion or around the difference of two proportions were done according to a binomial distribution using the Wilson Score method. We used the term sensitivity to refer to the proportion of accurately identified RADT result interpretations of positive and of invalid results; specificity referred to accurate RADT result interpretation of negative results.

Generalized linear mixed-effects models were fit to evaluate the association of the use of the Modified Quick Reference Guide with accurate RADT proficiency panel result interpretation. Outbreak business and participant were modelled as a random intercepts; participant age (in years), gender, RADT result type (positive; strong positive; negative; invalid) and an interaction term between RADT result type and instruction type were included as covariates (fixed effects).

Data were analyzed using R version 3.5.2 (R Core Team, Vienna, Austria). Statistical significance was assessed by using 2-tailed tests, with an α of 0.05.
